# Knowledge, attitude, and practice regarding hepatitis B virus among healthcare workers in Beira City, Mozambique

**DOI:** 10.1101/2025.05.20.25327983

**Authors:** Lúcia Vieira, Orvalho Augusto, Nédio Mabunda

## Abstract

**Background:** Hepatitis B is a disease transmitted through blood and other body fluids. In addition to vaccination, knowledge about the disease and appropriate practices help healthcare workers to prevent the disease. This study aimed to assess the knowledge, attitudes and practices of healthcare workers in Beira City, Mozambique.

**Methods:** This is a cross-sectional study, carried out in 4 health units in Beira City, Mozambique. It took place between June and August 2020. The sampling was systematically random, involving healthcare workers in direct contact with patients. A structured questionnaire was used to assess knowledge, attitudes, and practices.

**Results:** Of the 315 healthcare workers who took part in the study, the majority were nurses (125, 39.8%). The averages for correctly answered knowledge, attitude, and practice questions were 69.6%, 92.0%, and 54.8%, respectively. Physicians obtained the highest average percentage of correct items in all questions (85.3%) related to HBV knowledge and the nurse had highest attitude score (93.2%). Laboratory technicians were more correct about safe practices for HBV (60.1%). Workers in the medical department showed better knowledge of HBV compared to those working in the laboratory MR 0.84 (95% CI; 0.73 - 0.97), pediatrics MR 0.74 (95% CI; 0.62 – 0.89), outpatient consultations MR 0.89 (95% CI; 0.81 – 0.98), and other departments RM 0.80 (95% CI; 0.70 – 0.91. Those who had PCI training had better knowledge MR 0.88 (0.80 - 0.98) and practice 0.94 (0.91 - 0.98).

**Conclusion:** This study showed moderate knowledge and good attitudes about HBV among health workers. Practices regarding HBV were mediocre for all health workers. The data from this study highlight the need for training and the provision of materials and guidelines for the prevention and management of HBV as part of strategies to eliminate HBV as a public health problem.

## INTRODUCTION

Hepatitis B virus (HBV) is an important global public health problem. About one million new HBV infections occur daily and 254 million people are living with chronic hepatitis B (1). Among infected people worldwide, about 1.5 million people die annually from complications associated with HBV, like acute hepatitis, liver cirrhosis and hepatocellular carcinoma (HCC). In 2022, hepatitis B caused an estimated 1.1 million deaths (83% of all viral hepatitis deaths(1). In Mozambique)in 2019 about 2,370,020 people were living with chronic Hepatitis B and 736 died from the disease, 40 of which died from HCC associated with HBV. Currently, the mortality rate in the country is 2.49 people per 100,000 inhabitants per year(2,3).

Mozambique’s prevalence of HBV has been estimated to be over 8% which is considered to be a high level(4). Studies carried out locally with various population groups also show this high prevalence of HBV in Mozambique. In Youth of both genders aged 18–24 years a prevalence of 12.2% (5) was found. In persons who self-reported to have ever injected drugs 32.8% (6) was found and in untreated HIV-infected adults 9.1%(7). A study recently carried out for the first time among healthcare workers (HCWs) in the second largest city in Mozambique, showed an intermediate prevalence of 5.1%, with 27.9% of HCWs susceptible to the infection, 6.3% naturally immune and 60% immune by vaccination(8).

Developing countries such as Mozambique have limited resources to implement HBV elimination strategies(9). The childhood vaccination against HBV is part of the National Vaccination Program, occurring in the second, third and fourth month of life, however, vaccination for adults, high-risk groups or HCWs is not routinely available (10). Testing only occurs in people with HBV symptoms and during blood transfusions. Routine HBV testing for pregnant women during antenatal care is not available. The non-implementation of some HBV elimination strategies, associated with non-compliance with individual preventive measures, such as unprotected sex and multiple sexual partners, contribute to the high prevalence of HBV in the country(11).

HCWs are an important risk group for HBV infection. The factors associated with occupational infection in health workers are precarious working conditions, insufficient training and high workload (9). These risks can be mitigated with the implementation of some measures such as training in hospital infection control procedures, availability of personal protective equipment and timely vaccination(4). Studies show that preventive measures against HBV are not widely implemented in HCW, especially in developing countries. The vaccination rate among health professionals is low, the availability of resources for patient management is scarce and post-exposure prophylaxis is not fully effective (12,13).

Studies carried out in Africa about knowledge, attitudes and practices (KAP) show that those with a high level of education have better levels of knowledge about HBV than those with low levels of education. The balance between the knowledge and practices of the disease is important for a more effective protection. Those with lower levels of education have less knowledge about the HBV, mainly on transmission routes (14,15). Regarding practices, we find that the lack of work equipment, including adequate personal protective equipment (PPE), interferes with HCW protection. Most HCW are not vaccinated against HBV due to lack of opportunities to do so in their workplaces(16,17). The HBV vaccine is especially important for HCWs who are at constant risk of infection. Data for HBV prevalence and vaccination status in HCWs are not routinely collected in Mozambique. Furthermore, there are no published studies to determine the knowledge, attitudes, and practices. This study aims to evaluate the attitudes, knowledge, and practices of HCWs in relation to HBV, in four health facilities in the city of Beira.

## MATERIALS AND METHODS

### Study design, population, and area

This was a cross-sectional descriptive study, carried out in four health facilities (HF) in Beira City, Sofala province. This study was carried out in Beira Central Hospital (HCB) and in three other health facilities, namely, Ponta-Gêa, Nhaconjo and Chingussura. The HCB is a quaternary-level hospital, a reference hospital providing the highest level of care in the central region of the country, with about 900 beds for hospitalization and 69% of their workers being from Beira City. The hospital receives patients from the city of Beira with serious health conditions and patients transferred from health facilities in the three neighboring provinces. Of the HF selected, two were in an urban area (HCB and Ponta-gêa) and one was located in a peri-urban area (Nhaconjo and Chingussura).

The populations of this study were HCWs which had direct contact with patients (physicians, nurses, laboratory technicians, auxiliary staff, and others). The total study population was 1744.

Data collection for the study took place between data between June 9th and August 28th, 2020. The criteria for selecting the HF were the those with the highest volume and that there were no ongoing campaigns by isolated initiatives (Ex: by an NGO) for vaccination against hepatitis B in the last 5 years.

Inclusion criteria were HCWs who had contact with body fluids and handling sharp materials, was working at the time of data collection for the study, working in the health unit for at least 3 months and with sufficient physical and mental capacity to give free and informed consent.

### Sample size and sample selection

The sample size was estimated to be representative of the HCWs providing care and handling sharps in public healthcare facilities in Beira City. The formula for the sample size of a proportion with finite population correction was used through the OpenEpi web page (18) to detect a 50% proportion with a margin of error of 5% and at a 5% significance level among the total 1,744 HCWs. The final sample size was 315, which is 18.1% of HCWs in the city.

The sample was randomly selected with stratification by professional cadre (physicians, nurses, laboratory technicians, auxiliary staff and other providers with non-university degrees including dental and oral care, eye care, preventive medicine, instrumentalists, anesthesiology technicians, ophthalmology technicians, radiology technicians) maintaining the overall fraction of 18.1% (315/1,744). Within each strata a systematic selection was performed from a nominal list of providers. In case of non-consent, the next HCW on the list was called.

### Data collection

Data were collected using an electronic questionnaire inserted into a tablet and administered by an interviewer. Participants followed the questions through a physical form that was given before the interviews. The interviews were carried out in a room previously indicated in each sector. First, the team presented itself to HF directors, followed by the heads of the sectors, where they asked for a list of HCWs who were working in that sector and the indication of a private room, with no movement of people to carry out the interviews. The list was used to select the participants and after that it was safely discarded.

Participants were informed about the study as well as the location of the interviews by the head of their sector. The questionnaire was in Portuguese, containing closed questions and was divided into three parts. The first part consisted of sociodemographic questions, and the second, third and fourth parts contained questions about knowledge, attitudes and practices about HBV respectively. An ODK Collect application with the study questions was installed on each tablet. At the end of each working day, the data were sent to a server located at the institution. The tablets had a password-encrypted memory card.

### Statistical analysis

Data were collected in a Microsoft Excel spreadsheet (19) and then exported to R version 4.2.0(20) for analysis. Descriptive statistics were used, with frequencies and percentages for categorical variables, and means, standard deviations and quantiles for quantitative variables. A scoring system based on, respectively, 18, 8, and 9 correct answers was generated for knowledge, attitudes, and practices. We present the sum, the percentage and the average percentage of correct answers. For further analysis the average percentage of correct answers is used as the score. The analysis is stratified by professional cadre (physicians, nurses, laboratory technicians, auxiliary, and other cadres).

To study the association between health provider characteristics and the scores of knowledge, attitude and practices we conduct unadjusted and adjusted analysis. We use the mean ratio (MR) as a measure of association. The MR is a ratio of means from two groups, for example, the result of the division of mean score among males by the mean score among females. To produce the MR we use a generalized linear model (GLM) regression with Gaussian family, log-link and robust standard errors (21). For the adjusted analysis we include all collected health provider characteristics. In addition, to assess the associations with the 3 scores simultaneously we fit a generalized linear mixed model (GLMM) with the provider as the random-intercept.

### Ethical considerations

This study was approved by the National Committee of Bioethics for Health of Mozambique, with reference 412/CNBS/2020. Written informed consent was obtained from all healthcare workers before starting the interviews. The anonymity and confidentiality of the collected information was guaranteed.

## RESULTS

### Sociodemographic characteristics of study participants

Of the 315 healthcare workers who took part in the study, the majority were nurses (125, 39.8%) and the least represented group were laboratory technicians (26, 8.2%). In terms of gender, women were fewer in all professional categories, except for nurses and auxiliary staff, with (89, 71.2%) and (52, 63.4%) respectively, as shown in table 1. The 30–39 years old age group was the most represented (127, 40.3%), and the median age was 37.0 years (IQR, 31.5-46 years). Most of the participants (243, 77.1%) had more than 5 years of service.

**Table 1.**
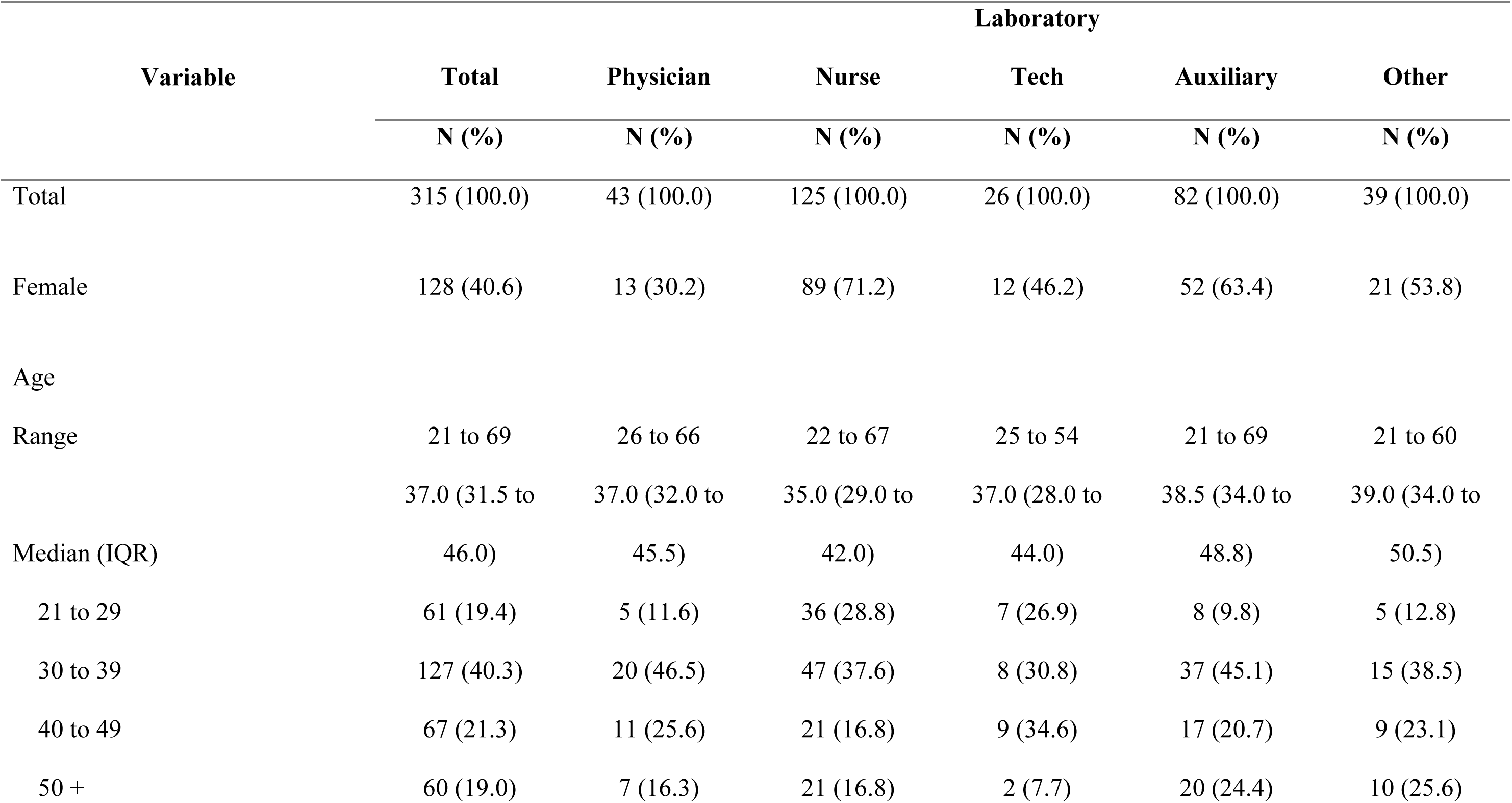
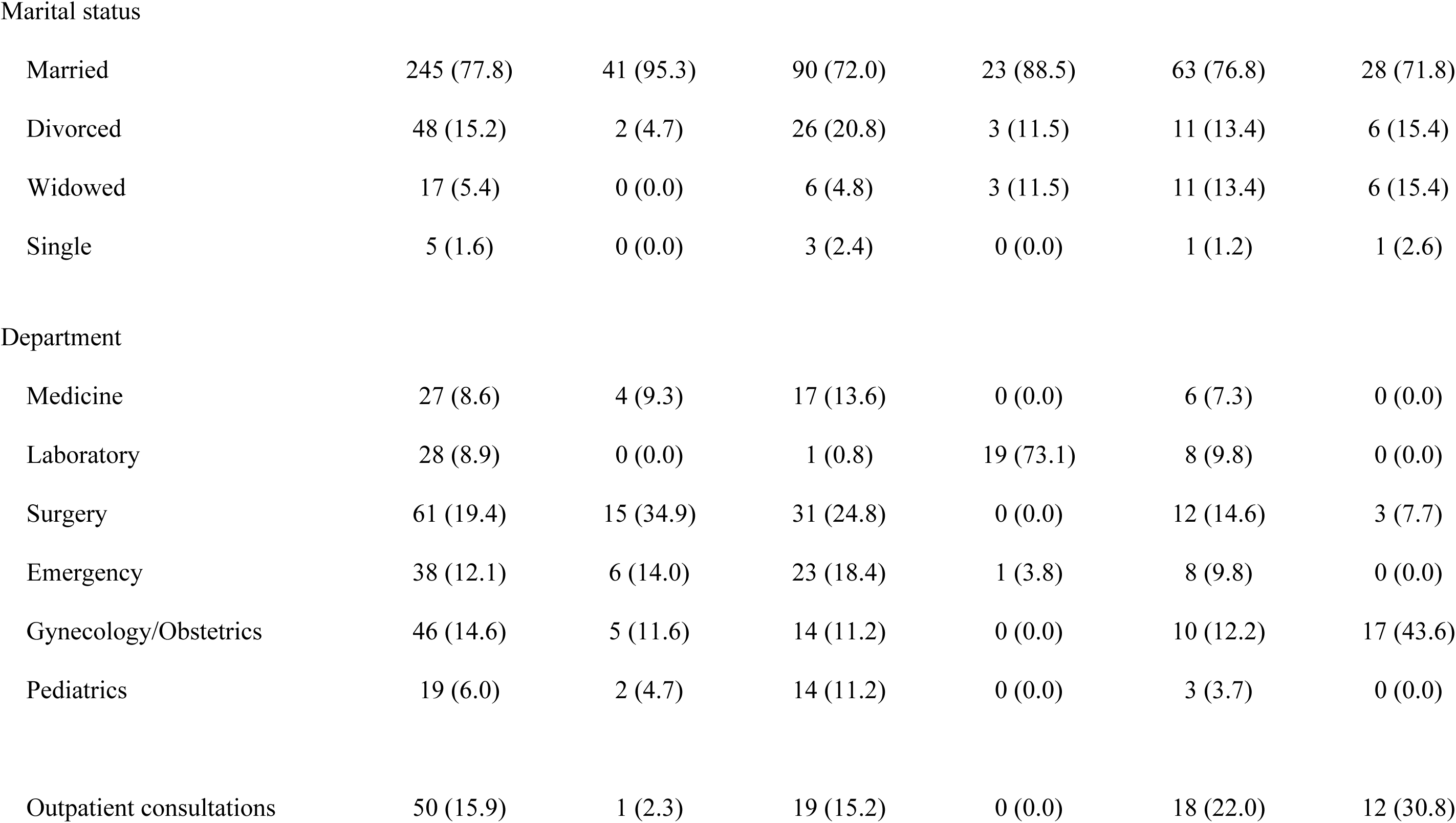
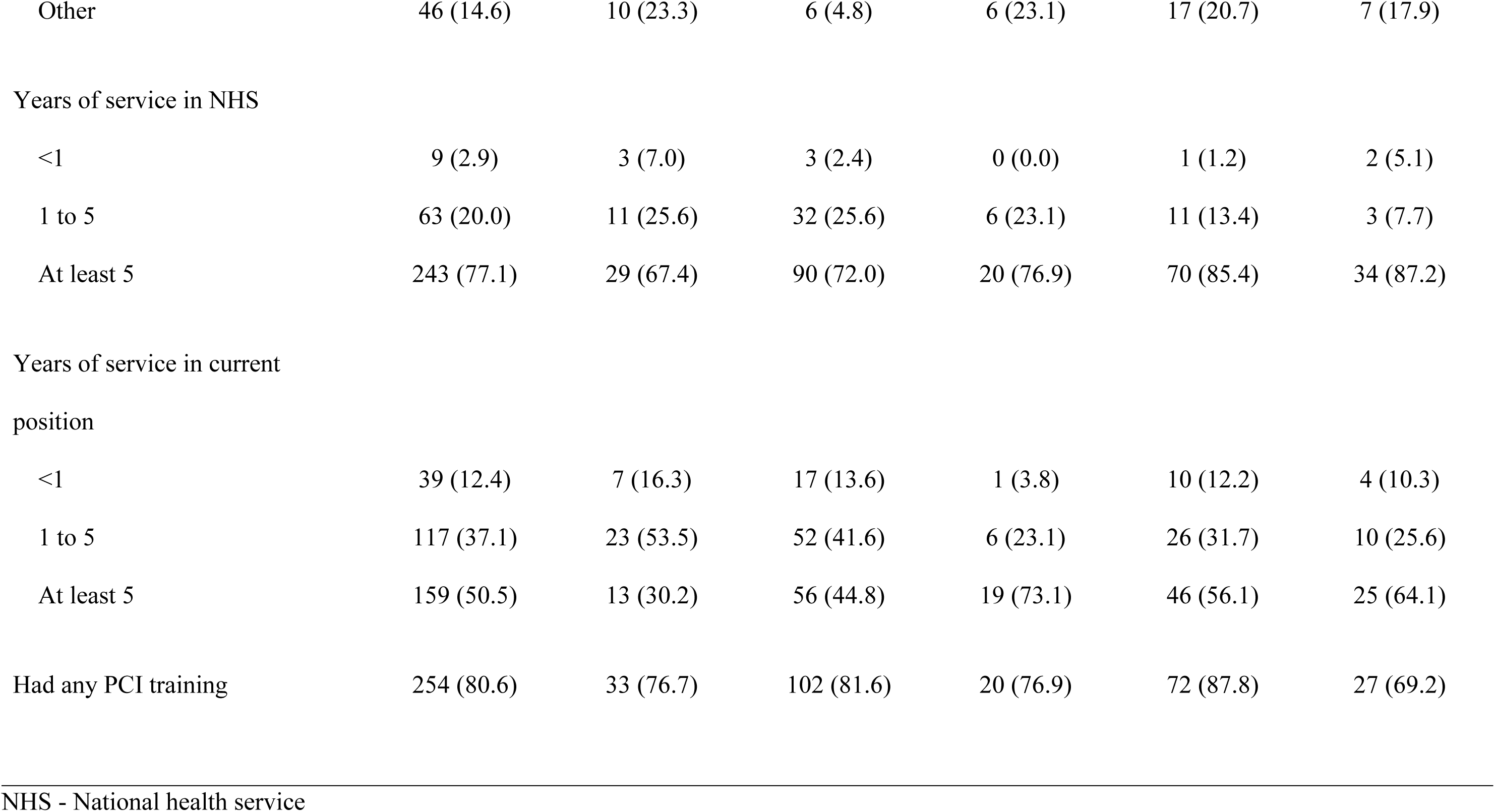
Sociodemographic characteristics of study participants.

### Assessment of knowledge and attitudes towards HBV

The assessment of the level of knowledge regarding HBV was measured through 18 questions. The physicians had the highest average percentage of correct items for all questions, 85.3% (IQR, 14.5; 17), while the auxiliary staff had the lowest average percentage of correct items, 56.4% (IQR, 7.3; 13), table 2. Except for physicians, the other professional categories scored lower on questions linked to symptoms of HBV infection. The question on the sexual transmission of HBV had the fewest correct answers.

**Table 2.**
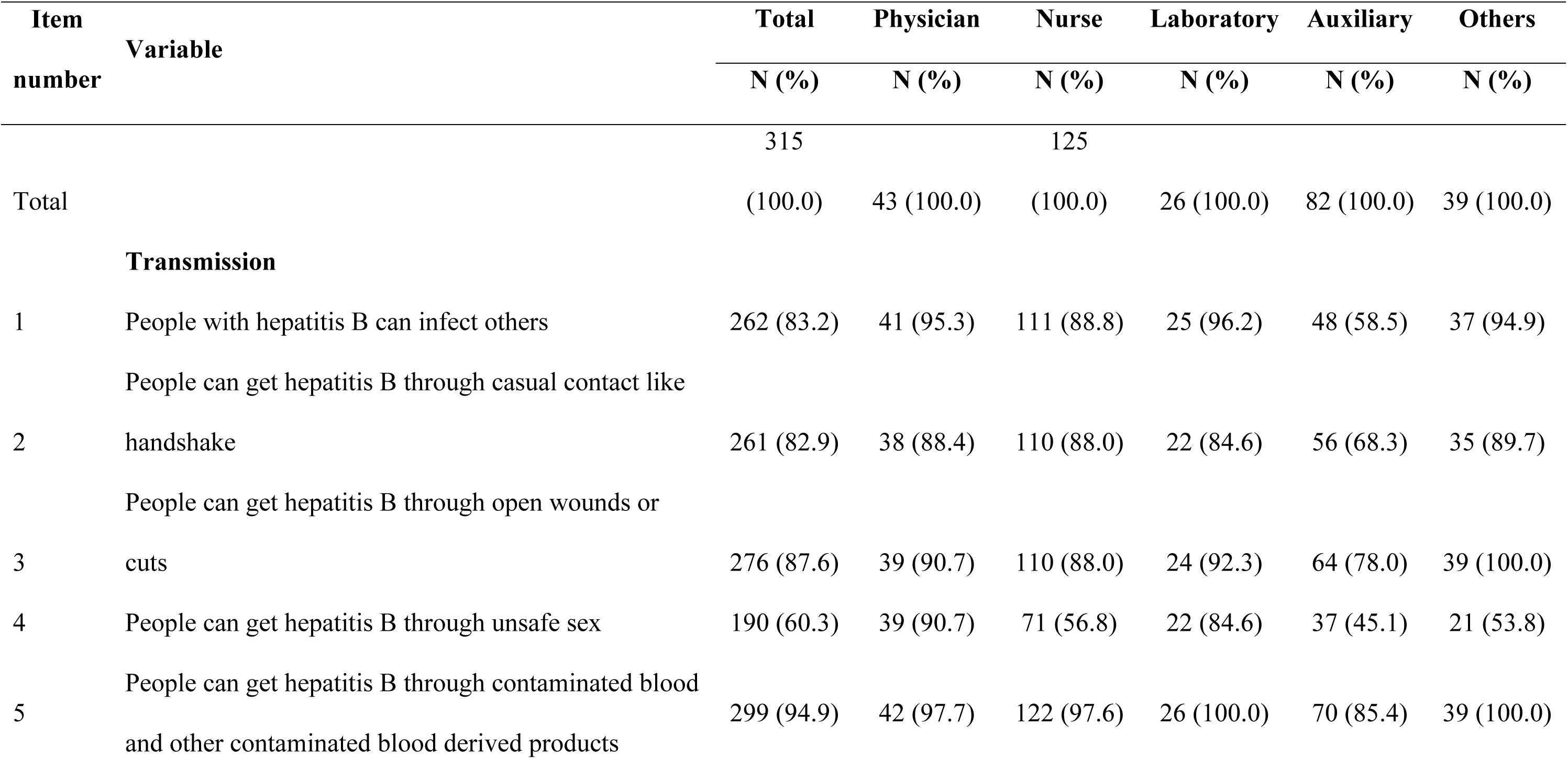
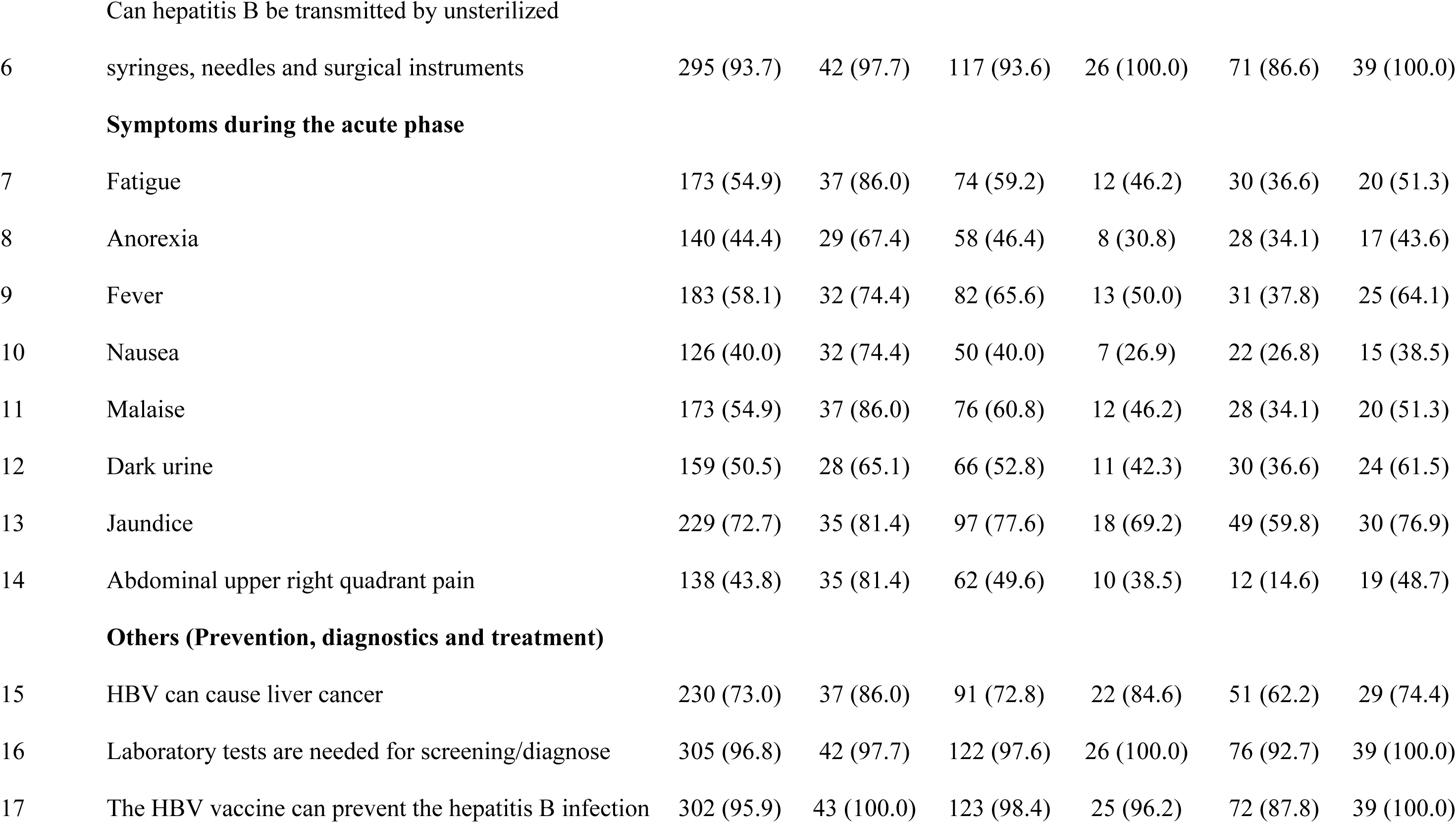
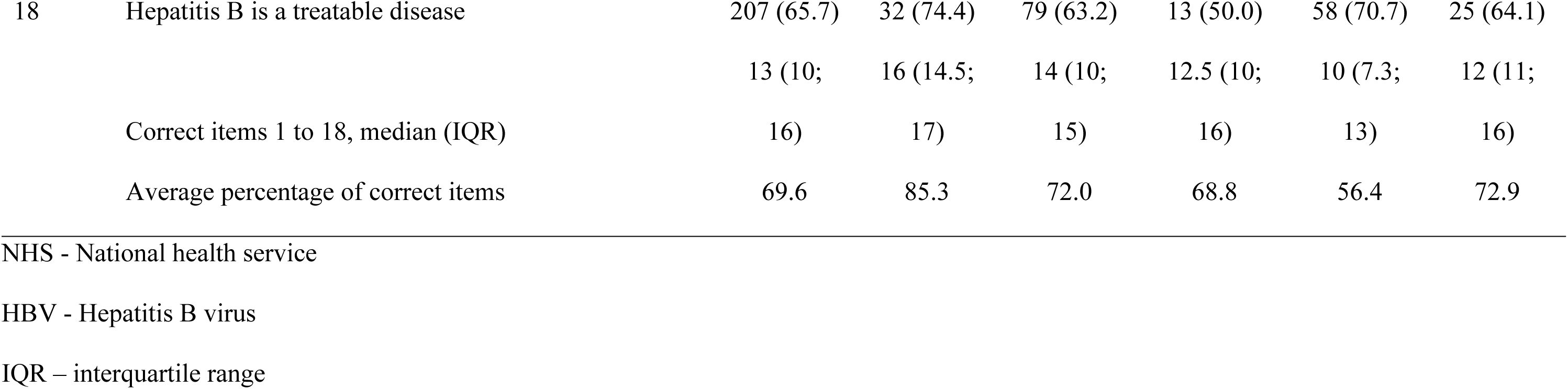
Knowledge towards HBV stratified by professional cadre.

The eight questions assessing attitudes towards HBV infection had an average percentage of correct attitudes of 92.0% (IQR, 7; 7.4). All professional categories had an average percentage of correct answers above 90%, table 3.

**Table 3.**
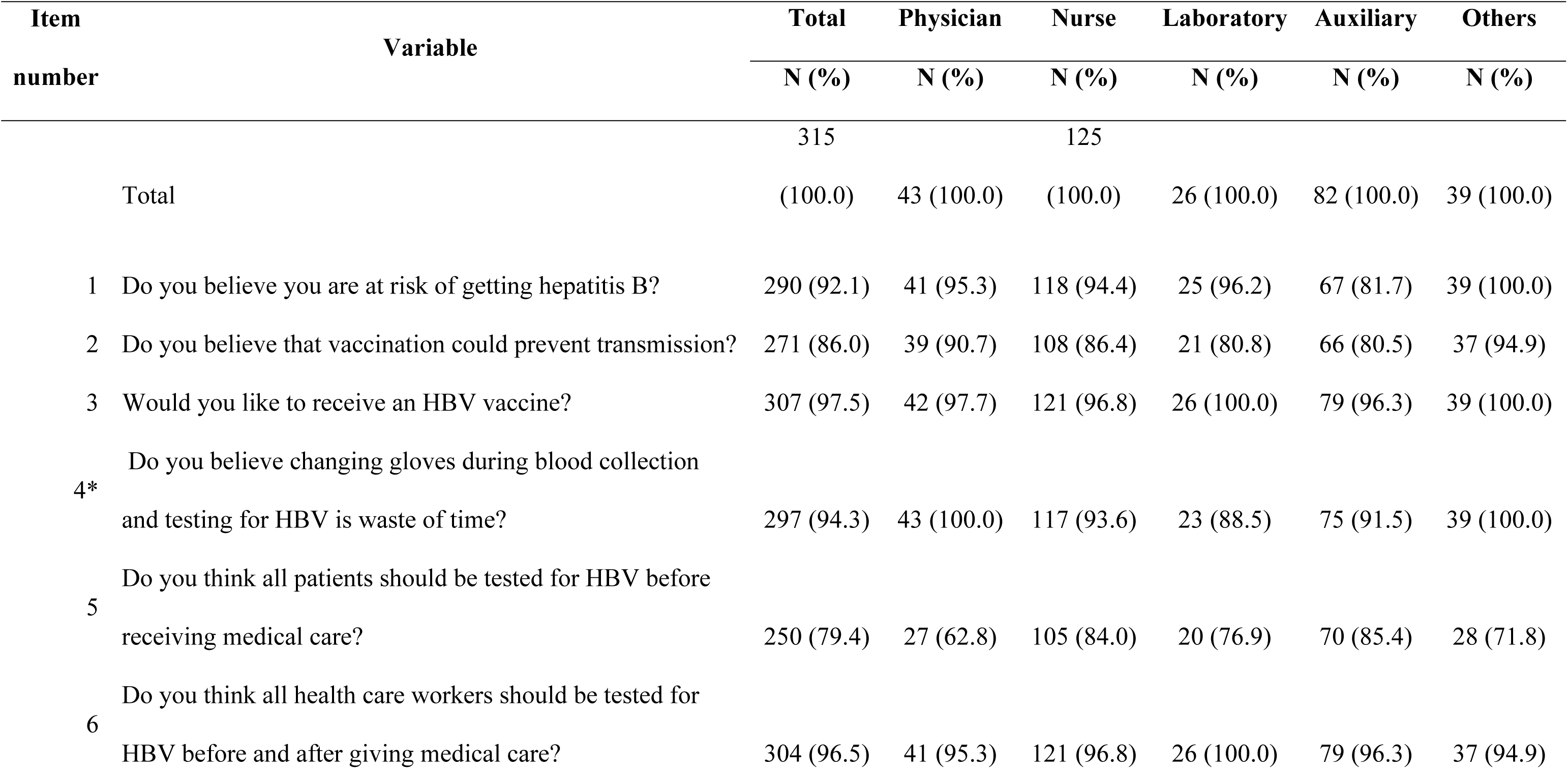
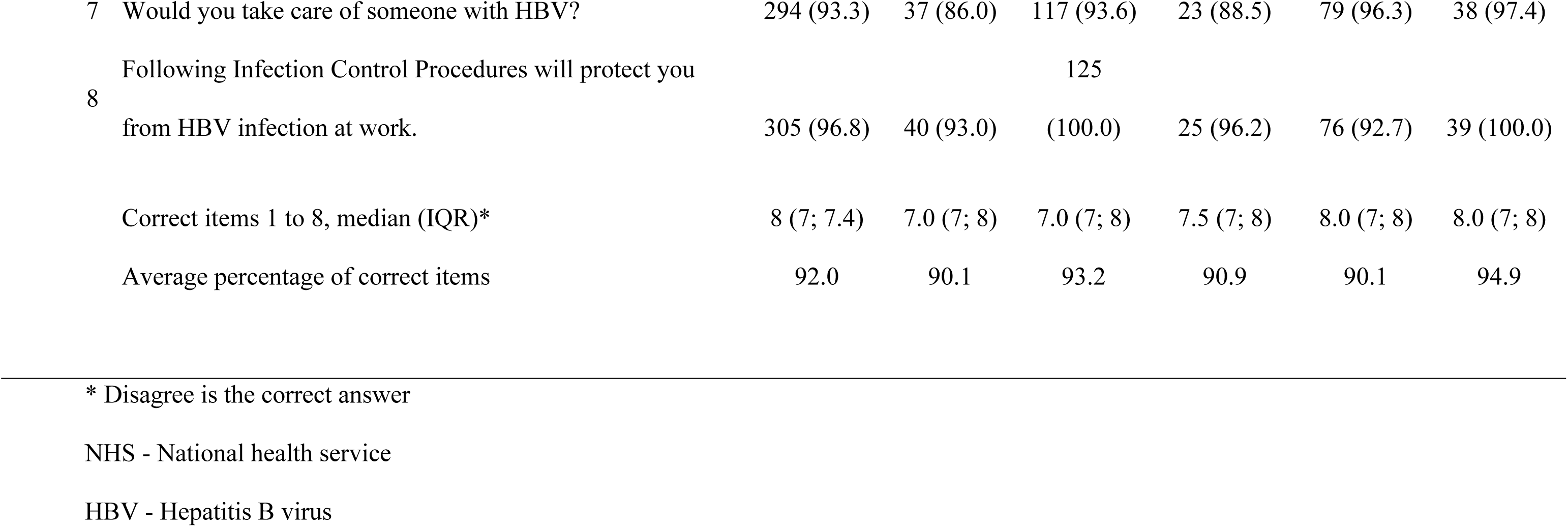
Attitudes towards HBV.

### Assessment of practice towards HBV

The overall average percentage of correct practices in relation to HBV was 50.3% (IQR, 3; 4), table 4. Laboratory technicians had the highest average percentage of correct practices with 56.8% (IQR, 3; 4). The use of personal protective equipment (PPE) before procedures involving body fluids and hand washing with soap and water were the practices with the highest scores, with 81.9% (258) and 86.0% (271) respectively.

**Table 4.** Safe practices regards HBV.

| Item<br>number | Variable | Total | Physician | Nurse | Laboratory | Auxiliary | Others |
| --- | --- | --- | --- | --- | --- | --- | --- |
|  |  | N (%) | N (%) | N (%) | N (%) | N (%) | N (%) |
|  |  | 315 |  | 125 |  |  |  |
|  | Total | (100.0) | 43 (100.0) | (100.0) | 26 (100.0) | 82 (100.0) | 39 (100.0) |
| 1 | Did test for HBV | 125 (39.7) | 21 (48.8) | 45 (36.0) | 19 (73.1) | 19 (23.2) | 21 (53.8) |
| 2 | Had HBV vaccine | 132 (41.9) | 26 (60.5) | 44 (35.2) | 19 (73.1) | 24 (29.3) | 19 (48.7) |
| 3 | Completed the HBV vaccine schedule | 43 (13.7) | 8 (18.6) | 10 (8.0) | 8 (30.8) | 8 (9.8) | 9 (23.1) |
| 4 | Change gloves between patients for blood collection | 239 (75.9) | 39 (90.7) | 96 (76.8) | 11 (42.3) | 59 (72.0) | 34 (87.2) |
| 5 | History of NSI or body fluids contact | 109 (34.6) | 18 (41.9) | 49 (39.2) | 7 (26.9) | 22 (26.8) | 13 (33.3) |
| 6 | Precautions should be taken with syringes after use | 121 (38.4) | 17 (39.5) | 62 (49.6) | 7 (26.9) | 21 (25.6) | 14 (35.9) |
| 7 | Wear PPE for medical procedures involving body fluids | 258 (81.9) | 30 (69.8) | 92 (73.6) | 25 (96.2) | 77 (93.9) | 34 (87.2) |
| 8 | Before disposing of infectious waste, is it chemically disinfected or autoclaved? | 127 (40.3) | 7 (16.3) | 45 (36.0) | 17 (65.4) | 39 (47.6) | 19 (48.7) |
| 9 | Washing hands with water & soap after NSI or body fluid contact | 271 (86.0) | 38 (88.4) | 115 (92.0) | 20 (76.9) | 64 (78.0) | 34 (87.2) |
|  | Correct items 1 to 9, median (IQR) | 4 (3; 4) | 4 (3; 4.5) | 4 (3; 4) | 4 (3; 4) | 4 (2; 4) | 4 (3; 5) |
|  | Average percentage of correct items | 50.3 | 52.7 | 49.6 | 56.8 | 45.1 | 56.1 |

NSI - Needle stick injury

### Association between demographic characteristics and KAP

Workers in the medical department showed better knowledge of HBV compared to those working in the laboratory MR 0.84 (95% CI; 0.73 - 0.97), pediatrics MR 0.74 (95% CI; 0.62 – 0.89), Outpatient consultations MR 0.89 (95% CI; 0.81 – 0.98), and other departments MR 0.80 (95% CI; 0.70 – 0.91), table 5 and 6.

**Table 5.**
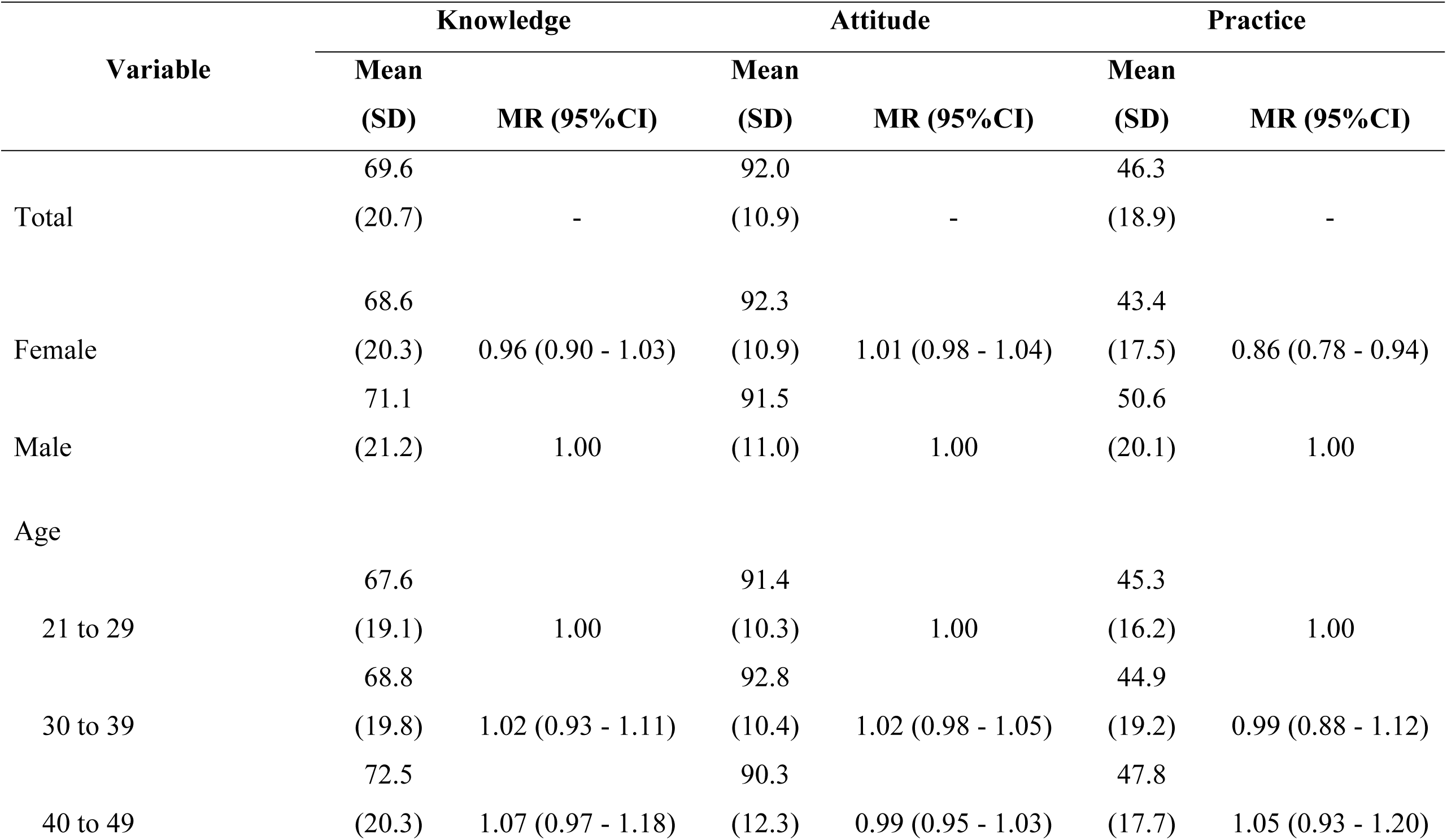
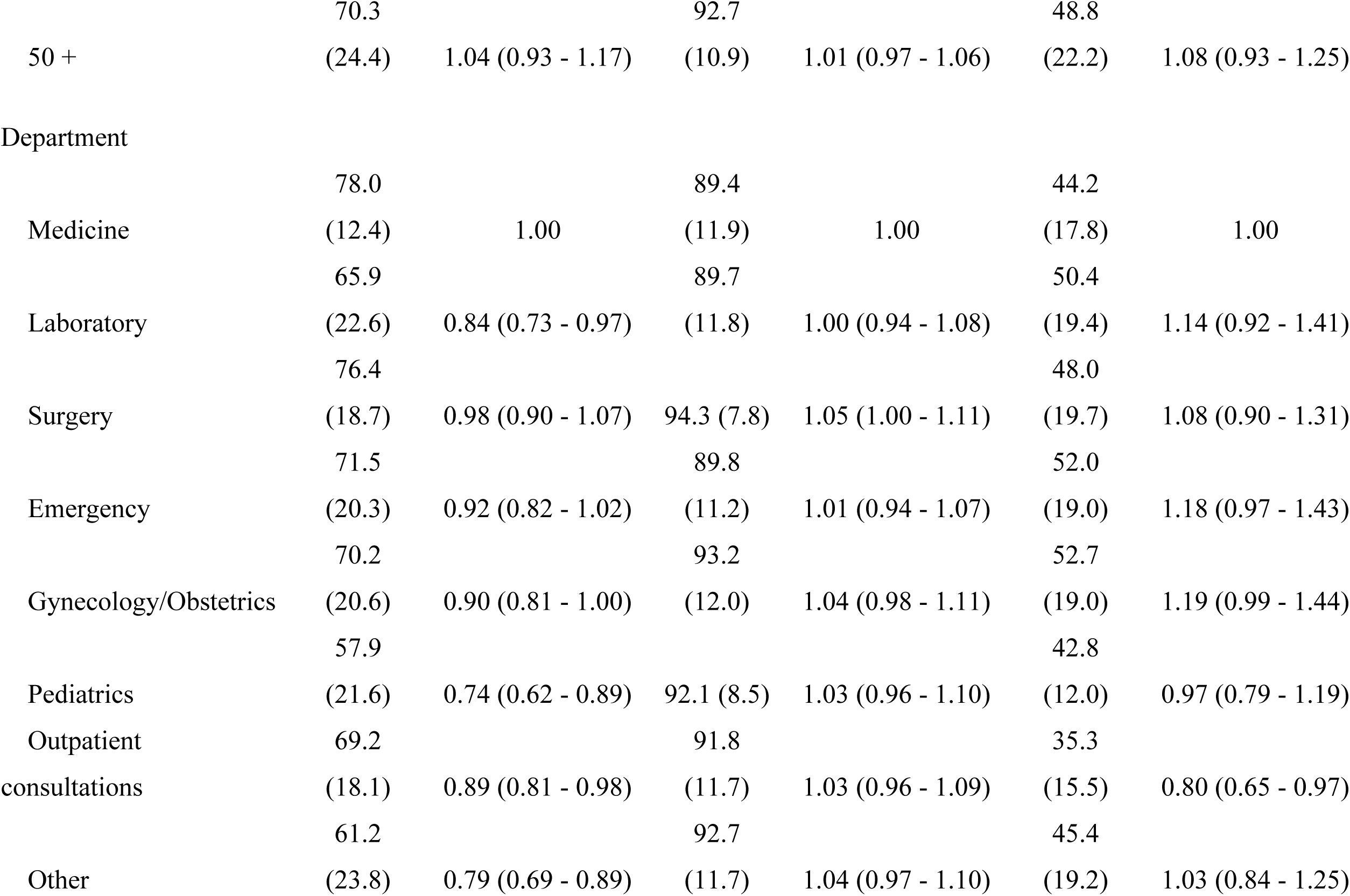
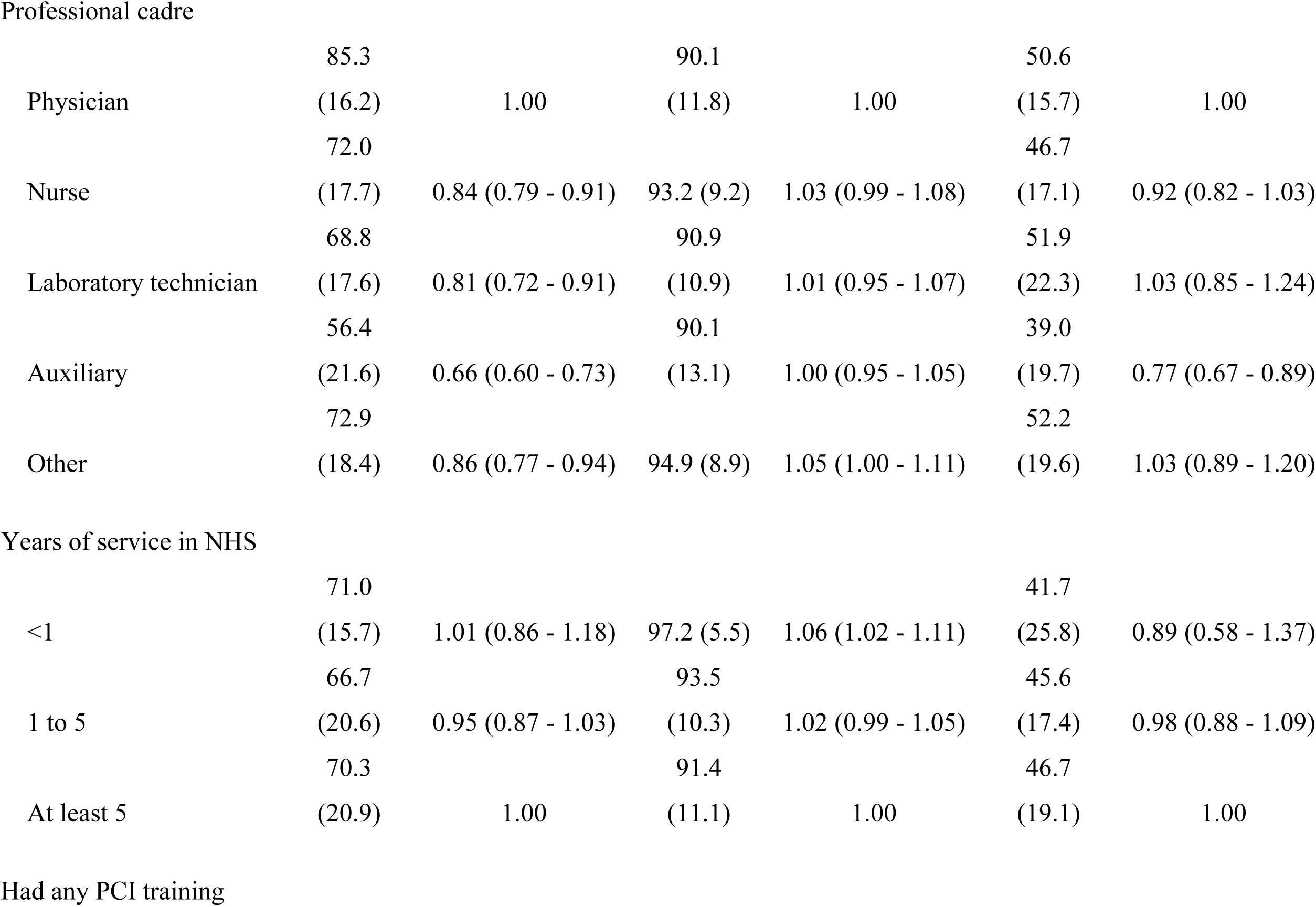
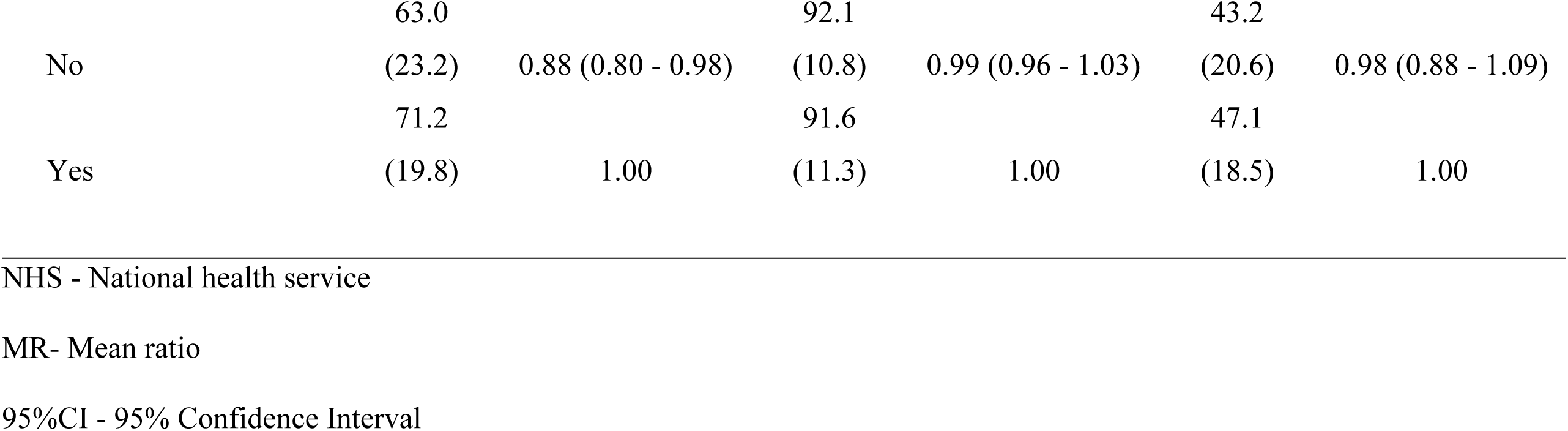
Mean average percentages scores, and crude associations between demographic characteristics and KAP scores.

| Variable | Knowledge |  | Attitude |  | Practice |  |
| --- | --- | --- | --- | --- | --- | --- |
|  | Mean<br>(SD) | MR (95%CI) | Mean<br>(SD) | MR (95%CI) | Mean<br>(SD) | MR (95%CI) |
| Total | 69.6<br>(20.7) | - | 92.0<br>(10.9) | - | 46.3<br>(18.9) | - |
| Female | 68.6<br>(20.3) | 0.96 (0.90 - 1.03) | 92.3<br>(10.9) | 1.01 (0.98 - 1.04) | 43.4<br>(17.5) | 0.86 (0.78 - 0.94) |
| Male | 71.1<br>(21.2) | 1.00 | 91.5<br>(11.0) | 1.00 | 50.6<br>(20.1) | 1.00 |
| Age |  |  |  |  |  |  |
| 21 to 29 | 67.6<br>(19.1) | 1.00 | 91.4<br>(10.3) | 1.00 | 45.3<br>(16.2) | 1.00 |
| 30 to 39 | 68.8<br>(19.8) | 1.02 (0.93 - 1.11) | 92.8<br>(10.4) | 1.02 (0.98 - 1.05) | 44.9<br>(19.2) | 0.99 (0.88 - 1.12) |
| 40 to 49 | 72.5<br>(20.3) | 1.07 (0.97 - 1.18) | 90.3<br>(12.3) | 0.99 (0.95 - 1.03) | 47.8<br>(17.7) | 1.05 (0.93 - 1.20) |
|  | 70.3 |  | 92.7 |  | 48.8 |  |
| 50 + | (24.4) | 1.04 (0.93 - 1.17) | (10.9) | 1.01 (0.97 - 1.06) | (22.2) | 1.08 (0.93 - 1.25) |
| Department |  |  |  |  |  |  |
|  | 78.0 |  | 89.4 |  | 44.2 |  |
| Medicine | (12.4) | 1.00 | (11.9) | 1.00 | (17.8) | 1.00 |
|  | 65.9 |  | 89.7 |  | 50.4 |  |
| Laboratory | (22.6) | 0.84 (0.73 - 0.97) | (11.8) | 1.00 (0.94 - 1.08) | (19.4) | 1.14 (0.92 - 1.41) |
|  | 76.4 |  |  |  | 48.0 |  |
| Surgery | (18.7) | 0.98 (0.90 - 1.07) | 94.3 (7.8) | 1.05 (1.00 - 1.11) | (19.7) | 1.08 (0.90 - 1.31) |
|  | 71.5 |  | 89.8 |  | 52.0 |  |
| Emergency | (20.3) | 0.92 (0.82 - 1.02) | (11.2) | 1.01 (0.94 - 1.07) | (19.0) | 1.18 (0.97 - 1.43) |
|  | 70.2 |  | 93.2 |  | 52.7 |  |
| Gynecology/Obstetrics | (20.6) | 0.90 (0.81 - 1.00) | (12.0) | 1.04 (0.98 - 1.11) | (19.0) | 1.19 (0.99 - 1.44) |
|  | 57.9 |  |  |  | 42.8 |  |
| Pediatrics | (21.6) | 0.74 (0.62 - 0.89) | 92.1 (8.5) | 1.03 (0.96 - 1.10) | (12.0) | 0.97 (0.79 - 1.19) |
| Outpatient | 69.2 |  | 91.8 |  | 35.3 |  |
| consultations | (18.1) | 0.89 (0.81 - 0.98) | (11.7) | 1.03 (0.96 - 1.09) | (15.5) | 0.80 (0.65 - 0.97) |
|  | 61.2 |  | 92.7 |  | 45.4 |  |
| Other | (23.8) | 0.79 (0.69 - 0.89) | (11.7) | 1.04 (0.97 - 1.10) | (19.2) | 1.03 (0.84 - 1.25) |
| Professional cadre |  |  |  |  |  |  |
|  | 85.3 |  | 90.1 |  | 50.6 |  |
| Physician | (16.2) | 1.00 | (11.8) | 1.00 | (15.7) | 1.00 |
|  | 72.0 |  |  |  | 46.7 |  |
| Nurse | (17.7) | 0.84 (0.79 - 0.91) | 93.2 (9.2) | 1.03 (0.99 - 1.08) | (17.1) | 0.92 (0.82 - 1.03) |
|  | 68.8 |  | 90.9 |  | 51.9 |  |
| Laboratory technician | (17.6) | 0.81 (0.72 - 0.91) | (10.9) | 1.01 (0.95 - 1.07) | (22.3) | 1.03 (0.85 - 1.24) |
|  | 56.4 |  | 90.1 |  | 39.0 |  |
| Auxiliary | (21.6) | 0.66 (0.60 - 0.73) | (13.1) | 1.00 (0.95 - 1.05) | (19.7) | 0.77 (0.67 - 0.89) |
|  | 72.9 |  |  |  | 52.2 |  |
| Other | (18.4) | 0.86 (0.77 - 0.94) | 94.9 (8.9) | 1.05 (1.00 - 1.11) | (19.6) | 1.03 (0.89 - 1.20) |
| Years of service in NHS |  |  |  |  |  |  |
|  | 71.0 |  |  |  | 41.7 |  |
| <1 | (15.7) | 1.01 (0.86 - 1.18) | 97.2 (5.5) | 1.06 (1.02 - 1.11) | (25.8) | 0.89 (0.58 - 1.37) |
|  | 66.7 |  | 93.5 |  | 45.6 |  |
| 1 to 5 | (20.6) | 0.95 (0.87 - 1.03) | (10.3) | 1.02 (0.99 - 1.05) | (17.4) | 0.98 (0.88 - 1.09) |
|  | 70.3 |  | 91.4 |  | 46.7 |  |
| At least 5 | (20.9) | 1.00 | (11.1) | 1.00 | (19.1) | 1.00 |
| Had any PCI training |  |  |  |  |  |  |
|  | 63.0 |  | 92.1 |  | 43.2 |  |
| No | (23.2) | 0.88 (0.80 - 0.98) | (10.8) | 0.99 (0.96 - 1.03) | (20.6) | 0.98 (0.88 - 1.09) |
|  | 71.2 |  | 91.6 |  | 47.1 |  |
| Yes | (19.8) | 1.00 | (11.3) | 1.00 | (18.5) | 1.00 |

NHS - National health service
MR- Mean ratio
95%CI - 95% Confidence Interval

As for the professional cadre, physicians showed better knowledge than other professional categories. With the adjustment, it was observed that physicians had better knowledge compared to nurses MR 0.83 (95% CI; 0.77 - 0.90), auxiliaries MR 0.65 (95% CI; 0.59 - 0.72), and others MR 0.86 (95% CI; 0.77 - 0.97). On the other hand, workers who had any PCI training had better knowledge than those who had no training, RM 0.88 (95% CI; 0.80 - 0.98).

Nurses MR 1.05 (95% CI; 1.00-1.10), and others MR 1.06 (95% CI; 1.01 - 1.12), had better attitudes compared to physicians, table 6. It was also observed that technicians with between 1 and 5 years’ experience MR 1.04 (95% CI; 1.00-1.18) and professionals with less than 1 year’s experience MR 1.07 (95% CI; 1.00-1.13) had a better attitude than technicians with more than five years’ experience in the national health service.

**Table 6.**
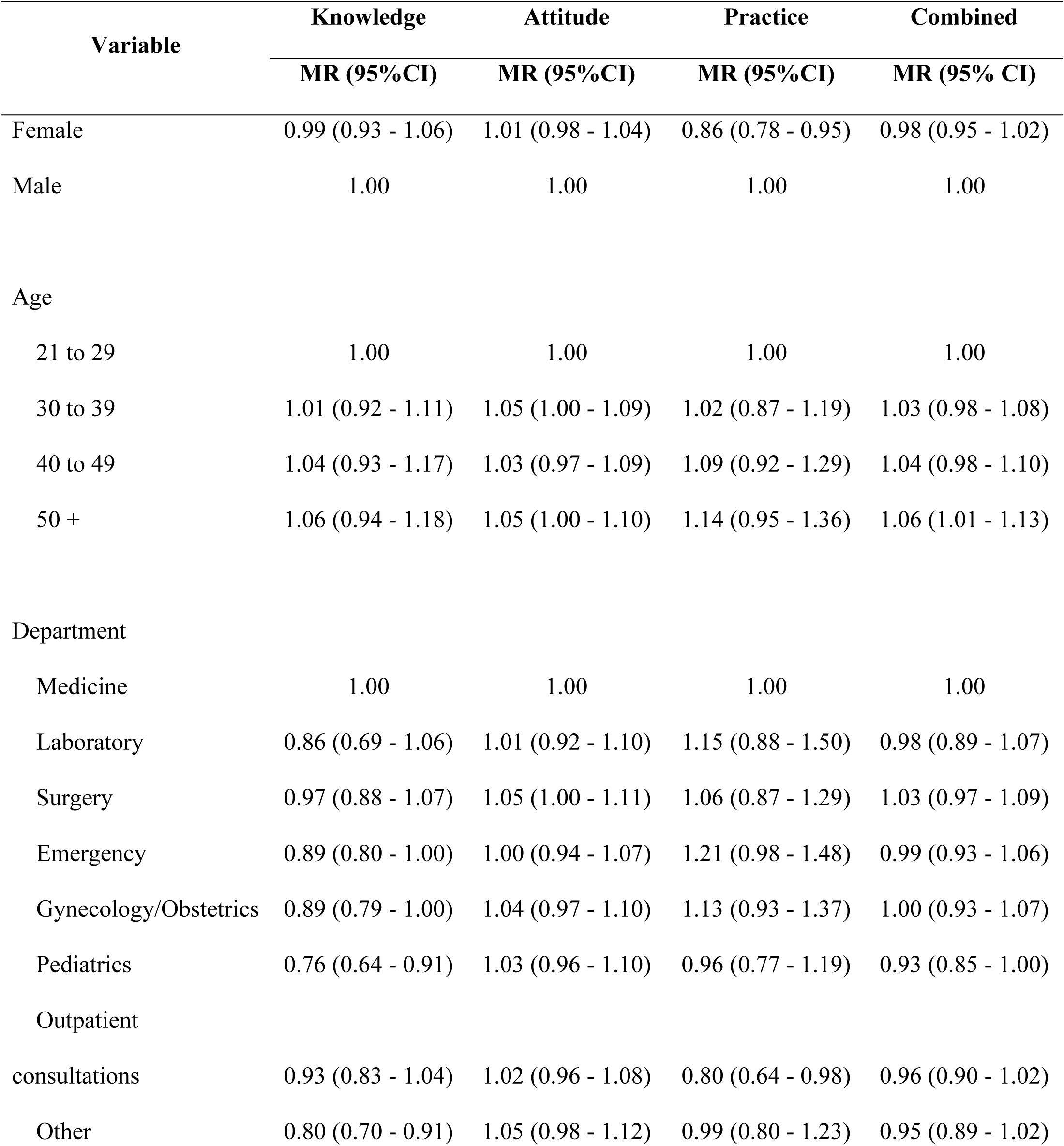
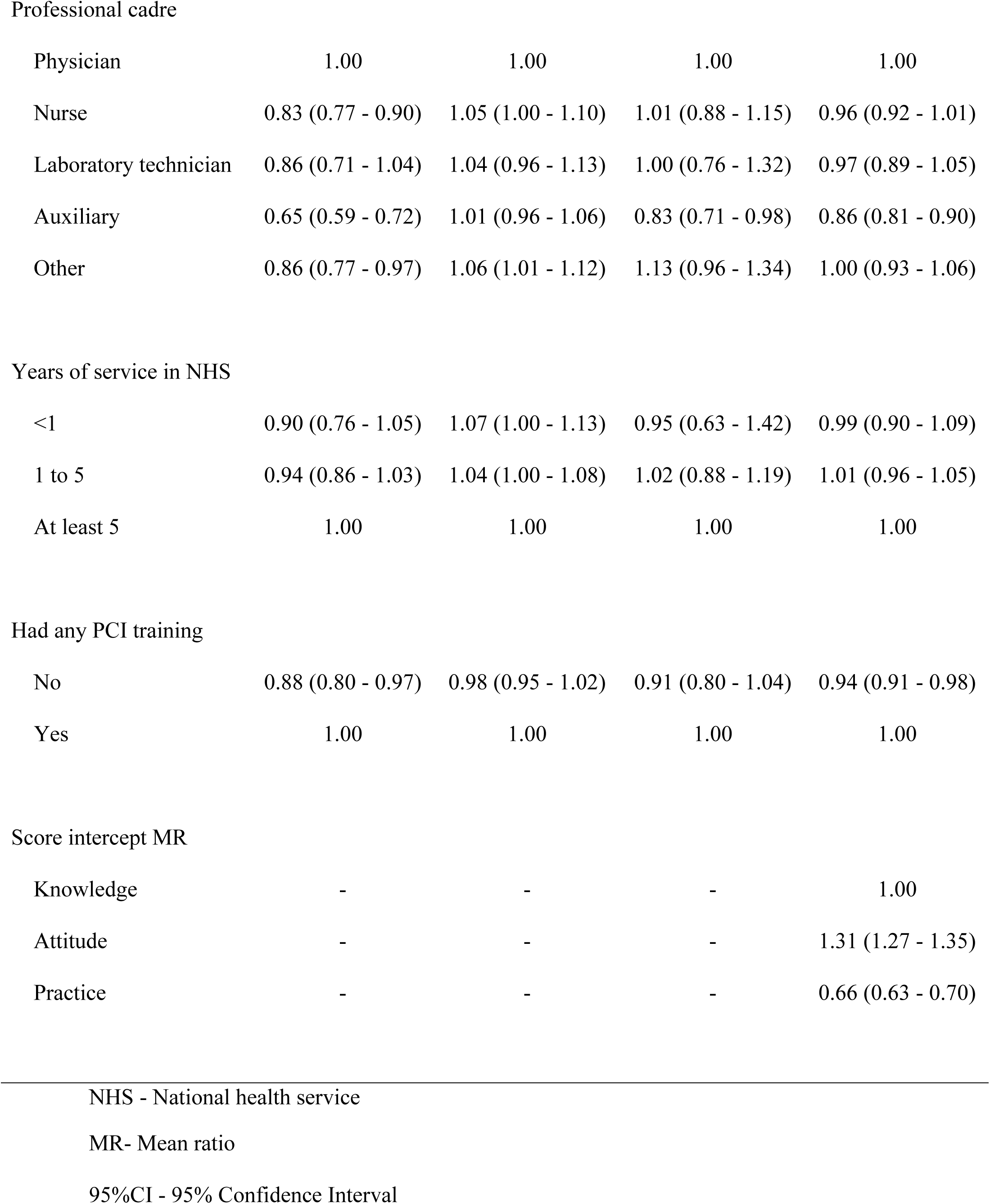
Adjusted associations between demographic characteristics and KAP scores.

| Variable | Knowledge | Attitude | Practice | Combined |
| --- | --- | --- | --- | --- |
|  | MR (95%CI) | MR (95%CI) | MR (95%CI) | MR (95% CI) |
| Female | 0.99 (0.93 - 1.06) | 1.01 (0.98 - 1.04) | 0.86 (0.78 - 0.95) | 0.98 (0.95 - 1.02) |
| Male | 1.00 | 1.00 | 1.00 | 1.00 |
| Age |  |  |  |  |
| 21 to 29 | 1.00 | 1.00 | 1.00 | 1.00 |
| 30 to 39 | 1.01 (0.92 - 1.11) | 1.05 (1.00 - 1.09) | 1.02 (0.87 - 1.19) | 1.03 (0.98 - 1.08) |
| 40 to 49 | 1.04 (0.93 - 1.17) | 1.03 (0.97 - 1.09) | 1.09 (0.92 - 1.29) | 1.04 (0.98 - 1.10) |
| 50 + | 1.06 (0.94 - 1.18) | 1.05 (1.00 - 1.10) | 1.14 (0.95 - 1.36) | 1.06 (1.01 - 1.13) |
| Department |  |  |  |  |
| Medicine | 1.00 | 1.00 | 1.00 | 1.00 |
| Laboratory | 0.86 (0.69 - 1.06) | 1.01 (0.92 - 1.10) | 1.15 (0.88 - 1.50) | 0.98 (0.89 - 1.07) |
| Surgery | 0.97 (0.88 - 1.07) | 1.05 (1.00 - 1.11) | 1.06 (0.87 - 1.29) | 1.03 (0.97 - 1.09) |
| Emergency | 0.89 (0.80 - 1.00) | 1.00 (0.94 - 1.07) | 1.21 (0.98 - 1.48) | 0.99 (0.93 - 1.06) |
| Gynecology/Obstetrics | 0.89 (0.79 - 1.00) | 1.04 (0.97 - 1.10) | 1.13 (0.93 - 1.37) | 1.00 (0.93 - 1.07) |
| Pediatrics | 0.76 (0.64 - 0.91) | 1.03 (0.96 - 1.10) | 0.96 (0.77 - 1.19) | 0.93 (0.85 - 1.00) |
| Outpatient |  |  |  |  |
| consultations | 0.93 (0.83 - 1.04) | 1.02 (0.96 - 1.08) | 0.80 (0.64 - 0.98) | 0.96 (0.90 - 1.02) |
| Other | 0.80 (0.70 - 0.91) | 1.05 (0.98 - 1.12) | 0.99 (0.80 - 1.23) | 0.95 (0.89 - 1.02) |
| Professional cadre |  |  |  |  |
| Physician | 1.00 | 1.00 | 1.00 | 1.00 |
| Nurse | 0.83 (0.77 - 0.90) | 1.05 (1.00 - 1.10) | 1.01 (0.88 - 1.15) | 0.96 (0.92 - 1.01) |
| Laboratory technician | 0.86 (0.71 - 1.04) | 1.04 (0.96 - 1.13) | 1.00 (0.76 - 1.32) | 0.97 (0.89 - 1.05) |
| Auxiliary | 0.65 (0.59 - 0.72) | 1.01 (0.96 - 1.06) | 0.83 (0.71 - 0.98) | 0.86 (0.81 - 0.90) |
| Other | 0.86 (0.77 - 0.97) | 1.06 (1.01 - 1.12) | 1.13 (0.96 - 1.34) | 1.00 (0.93 - 1.06) |
| Years of service in NHS |  |  |  |  |
| <1 | 0.90 (0.76 - 1.05) | 1.07 (1.00 - 1.13) | 0.95 (0.63 - 1.42) | 0.99 (0.90 - 1.09) |
| 1 to 5 | 0.94 (0.86 - 1.03) | 1.04 (1.00 - 1.08) | 1.02 (0.88 - 1.19) | 1.01 (0.96 - 1.05) |
| At least 5 | 1.00 | 1.00 | 1.00 | 1.00 |
| Had any PCI training |  |  |  |  |
| No | 0.88 (0.80 - 0.97) | 0.98 (0.95 - 1.02) | 0.91 (0.80 - 1.04) | 0.94 (0.91 - 0.98) |
| Yes | 1.00 | 1.00 | 1.00 | 1.00 |
| Score intercept MR |  |  |  |  |
| Knowledge | - | - | - | 1.00 |
| Attitude | - | - | - | 1.31 (1.27 - 1.35) |
| Practice | - | - | - | 0.66 (0.63 - 0.70) |

MR- Mean ratio
95%CI - 95% Confidence Interval

Technicians working in the Medicine Department had better practice than those working in the Outpatient consultations Department, MR 0.80 (95% CI; 0.65-0.97), table 6. On the other hand, physicians had better pratices than assistants, MR 0.83 (95% CI; 0.71 - 0.98).

## DISCUSSION

General knowledge of HBV among healthcare workers was moderate (69.6%), with physicians having the highest percentage of correct answers (85.3%). The results are similar to other studies carried out in Ethiopia (73.9%)(22), Cameroon (67.6%)(14), India (96.7%)(23), and Palestine (76.5%)(24). Knowledge levels below what we observed were reported in Democratic Republic of the Congo (33.2%)(13), Vietnam (59.5%)(25), and Uganda (50.8%)(26). The differences in results may be related to the level of implementation of HBV prevention, diagnosis, and treatment strategies in the countries, as well as the implementation of training after graduation and in-service training. The diversity and academic level of the healthcare workers in the different studies may also have influenced the results.

In general, there is low knowledge of HBV symptoms and transmission methods among non-physician healthcare workers. These results are similar to studies carried out in others African and Asian countries (23)(13)(14).

Better knowledge of HBV on the part of healthcare workers allows for clearer information for patients and the community in general, contributing to the prevention and control of HBV. The integration of HBV topics into HIV training could improve knowledge about HBV (15)(25).

Healthcare workers generally had a positive attitude towards HBV (92%). Although studies have reported that attitudes towards HBV are associated with knowledge of the disease (27) (23)(28), we believe that the results we found reflect general knowledge of similar diseases with greater awareness, such as HIV.

The practices regarding HBV were mediocre among healthcare workers (54.8%). Among the practices assessed, hand washing after any procedure involving contact with patients or body fluids was the most common among healthcare workers (86%). This may be related to the period of the study. In 2020, during the COVID-19 pandemic, there was instruction, recommendation, and a lot of information about handwashing to prevent COVID-19(29). The low level of practice among health workers about HBV may be related to several aspects: 1) Low implementation of chronic viral hepatitis elimination strategies in the country. Vaccination against HBV among health workers, for example, is not routine in Mozambique(8). In this study, less than half (41.9%) of health workers reported that they had been vaccinated against HBV and only 13.7% reported having had all three doses. Our previous results in this same population showed that a third of healthcare workers had no immunity to HBV(8). On the other hand, HBV testing in Mozambique is routine only in blood banks and is currently being implemented in the antenatal consultation; 2) Limited provision of collective and personal protection equipment. The SARA (national health inventory) observed that the national average availability of the necessary elements for infection control was 66%, with the majority of health facilities having limitations(30); 3) Lack of guidelines or poor dissemination of them. Continuous training for health workers ensures up-to-date and safe practices(15)(25). On the other hand, the availability of manuals, guidelines and other material allows for continuous consultation and self-training. Muchangos et al. observed the absence or rarity of normative documents related to the prevention, diagnosis and treatment of HIV/HBV co-infection in Mozambique(31).

Our study evaluates KAP in health workers in Mozambique for the first time. The training of health workers, their workplace, PCI training and professional experience are all aspects associated with KAP and should be taken into account in strategies to improve knowledge and practices in relation to HBV. However, some limitations must be taken into account when interpreting the results: 1) Possibility of selection bias, given the absence of health workers due to individual or service factors and refusal to participate for various reasons; 2) Participants may have suffered a memory bias due to the fact that the information provided about the exposure and vaccination event took place several years ago; 3) This study did not explore in more depth the reasons for non-vaccination against HBV and the period in which health workers were vaccinated. There may be cultural or other factors that limited vaccination; 4) This study may not be representative of other parts of the country because it was conducted in an urban area.

## CONCLUSION

Moderate knowledge and good attitudes about HBV were observed among health workers, with variation between the different categories of workers. Practices about HBV were mediocre for all health workers. The data from this study highlight the need for training and the provision of materials and guidelines for the prevention and management of HBV as part of strategies to eliminate HBV as a public health problem.

## SUPPORTING INFORMATION

**S1 File. Questionnaire for participants.**

**S2 File. Study data set.**

## Data Availability

All relevant data are within the manuscript and its Supporting Information files

## ACKNOWLEDGMENTS

We would like to gratefully acknowledge all the healthcare workers who participated in the study and Beira City health authorities for their collaboration. We extend thanks to Instituto Nacional de Saúde, Delegação Provincial de Sofala, especially Absalão Zumba, Ana Duaja, Angélica Sotomane and Falume Chale for their unconditional support in data collection and management. We also thank Dr. Sandy McGunegill for editing the English.

